# Sex hormones, obesity, and risk of cholecystectomy in men and women: a population-based prospective study and mediation analysis

**DOI:** 10.1101/2024.02.19.24303068

**Authors:** Jie-Qiong Lyu, Wei Jiang, Yi-Ping Jia, Meng-Yuan Miao, Jia-Min Wang, Hao-Wei Tao, Miao Zhao, Yong-Fei Hua, Guo-Chong Chen

## Abstract

**Background:** Obesity affects hormone metabolisms and contributes to gallstone disease more strongly in women than in men. This study assessed the sex-specific associations between serum levels of sex hormone-binding globulin (SHBG) and testosterone and risk of cholecystectomy, and their mediation role in the obesity-cholecystectomy association.

**Methods:** Included were 176,909 men and 160,147 women from the UK Biobank. Serum SHBG and total testosterone were measured by immunoassay. Incident cases of cholecystectomy for gallstone disease were identified using hospital inpatient records. Multivariable Cox proportional hazards models were used to calculate hazard ratios (HR) with 95% confidence interval (CI) of cholecystectomy associated with the serum hormones. A mediation analysis was performed to estimate the proportion of the obesity-cholecystectomy association potentially mediated by the two sex hormones.

**Results:** A total of 2877 men and 4607 women underwent cholecystectomies during the follow-up. Regardless of sex, higher levels of SHBG were associated with a lower risk of cholecystectomy. The HRs of cholecystectomy comparing the highest with the lowest quartiles of SHBG were 0.68 (95% CI: 0.59-0.77) in men (P-trend <0.001) and 0.39 (95% CI: 0.36-0.53) in women (P-trend <0.001). Higher levels of testosterone were associated with a higher risk of cholecystectomy in women (HR_Q4 vs. Q1_ = 1.28; 95% CI: 1.18-1.39; P-trend <0.001) but not in men (P-trend = 0.12). In women, it was estimated that 14.71% and 2.74% of the association between obesity and cholecystectomy was significantly medicated by SHBG and testosterone, respectively.

**Conclusions:** SHBG levels are inversely associated with risk of cholecystectomy in both sexes, whereas higher testosterone levels are associated with a higher risk of cholecystectomy only in women. Both hormones mediate the obesity-cholecystectomy association in women.

## Introduction

Gallstone disease is prevalent in developed countries, with an occurrence rate ranging from 10% to 15% in the US (1) and 15% in the UK (2). While most gallstones remain asymptomatic, complications such as cholecystitis, abdominal pain, cholangitis, and pancreatitis, occur in 20% of patients with gallstones (3). Such symptomatic gallstones often require surgical removal of the gallbladder through a surgical procedure, namely cholecystectomy, which is the standard treatment in such cases (4). As a frequently performed surgical procedure for gallstone disease, cholecystectomy contributes significantly to hospital admissions, healthcare costs, and patient morbidity (5, 6).

As compared with men, women have a considerably higher risk of gallstone disease (10), and seven in ten cholecystectomies are performed in women, potentially suggesting a role for sex hormones (11). Understanding factors that determine the sex disparity in gallstone formation may represent an opportunity for discovering novel risk factors for cholelithiasis. To this point, results from randomized controlled trial have shown that menopausal hormone replacement therapy (HRT) increase the risk for any gallbladder disease and cholecystectomy (12). Evidence from animal studies also supports the involvement of sex hormones in the development of gallstones (13, 14). A few epidemiological studies have examined the associations between SHBG or testosterone levels and risk of gallstone diseases, but the findings are inconsistent (15–18).

In addition, as a major risk factor for gallstone disease and a hormone-affecting factor, obesity increases the risk of gallstone disease more strongly in women than in men (19), although the underlying mechanisms for such sex-specific associations are unknown. Previous studies have suggested that the associations between obesity and sex-specific health outcomes may be partially mediated by sex hormones (20, 21). Whether and to what extent the association between obesity and the development of cholecystectomy could be mediated by circulating levels of SHBG or testosterone have not been assessed in previous studies.

In the present study, using data from a large population-based prospective study (UK Biobank), we investigated sex-specific associations between serum levels of SHBG and testosterone and the long-term risk of cholecystectomy. We also performed a mediation analysis to estimate the proportion of the obesity-cholecystectomy association potentially mediated by the two sex hormones.

## Methods

### Study Design and Population

The UK biobank is a large prospective cohort study that aims to investigate the primary causes of various chronic diseases, including genetic, environmental, and lifestyle factors (22). Details of the study protocol are available online (https://www.ukbiobank.ac.uk/media/gnkeyh2q/study-rationale.pdf). In brief, approximately 500,000 men and women aged 37 to 73 years were recruited between 2006 and 2010. Participants were invited to attend one of the 22 assessment centers located throughout England, Wales and Scotland where they underwent questionnaire surveys, brief interviews, and physical measurements. The UK Biobank study was approved by the research ethics committee (REC reference for UK Biobank 11/NW/0382) and all participants provided informed consent at recruitment.

Participants with a history of cholecystectomy (n = 8851), disorders of gallbladder, biliary tract, or pancreas (n = 10,709), cancer (except non-melanoma skin cancer; n = 43,562) at recruitment were excluded. We also excluded individuals who developed cancer in specific organs (the duodenum, liver, gallbladder, biliary ducts, or pancreas) (n = 1,968) or received liver transplantation (n = 48) during follow-up. Further excluded were participants without measurements of SHBG or testosterone (n = 96,958) and those who had the top 0.5% SHBG or testosterone levels in men or in women (n = 3259). Consequently, a total of 337,056 individuals (176,909 men and 160,147 women) were included for analysis (**Supplementary** Figure 1).

### Blood Sample Collection and Laboratory Assays

During recruitment, blood samples were collected from all participants at assessment centers and stored at a temperature of –80°C. Serum SHBG levels were measured using a two-step sandwich chemiluminescent immunoassay (DXI 800, Beckman Coulter, UK). Serum testosterone was assayed using a competitive binding chemiluminescent immunoassay (DXI 800, Beckman Coulter, UK). Comprehensive information regarding the assay techniques and quality assurance procedures can be accessed online (https://biobank.ndph.ox.ac.uk/showcase/showcase/docs/serum_biochemistry.pdf).

### Outcome Ascertainment

The outcome of interest in this study was the occurrence of cholecystectomy during the follow-up period. We chose cholecystectomy as the primary outcome because it is more clinically relevant than asymptomatic gallstones (23). The procedures of cholecystectomy were identified using hospital inpatient records obtained from the Hospital Episode Statistics for England, Scottish Morbidity Record data for Scotland, and the Patient Episode Database for Wales. Incident cases of cholecystectomy and the corresponding date of the procedures were identified using UK Biobank field IDs 41200 (operative procedures-main) and 41260 (date of first operative procedure-main).

### Assessment of Covariates

Information on sociodemographic factors, reproductive and medical histories, lifestyle behaviors, and medication use was collected at baseline by touchscreen questionnaires and nurse-led interviews. The Townsend deprivation index was derived by combining four census variables (unemployment, non-car ownership, non-home ownership, and household overcrowding). BMI was calculated based on measured weight and height (kg/m²). Physical activity was assessed at baseline using the self-reported short-form International Physical Activity Questionnaire and the data were summarized and reported in MET-h per week. Diabetes was defined as a self-reported physician’s diagnosis or antidiabetic medication use, or an HbA1c level of ≥6.5%.

### Statistical Analysis

Baseline characteristics of the participants were described by sex-specific quartiles of SHBG, for men and women separately. Continuous variables were presented as mean ± standard deviation (SD) and categorical variables as percentage. Spearman correlations were calculated among SHGB, testosterone, age, and BMI.

Cox proportional hazards models were employed to estimate hazard ratios (HRs) and 95% confidence intervals (CIs) of cholecystectomy across the sex-specific quartiles of SHBG or testosterone, using the lowest quartile of the analyzed sex hormone as the reference. We also modelled the sex hormones as continuous variables and estimated HRs (95%) of cholecystectomy for each one additional SD increment in SHBG or testosterone. Person-time of follow-up was calculated as the duration from date of baseline evaluation until the date of performing cholecystectomy, date of death or withdrawal from the study, or date of the most recent follow-up, whichever occurred first. Four Cox models were constructed to incrementally adjust for potential confounders.

Model 1 was adjusted for age (y), ethnicity (White, Asian or Asian British, Black or Black British, mixed ethnicities), and Townsend deprivation index. Model 2 was adjusted for the covariates in model 1 and was further adjusted for smoking status (never, former, current), drinking status (never, former, current), total physical activity (MET-h/week), diabetes (yes, no), and for women, menopausal status (yes, no) and HRT (yes, no). The full model (model 3) was adjusted for all the above listed covariates and was further adjusted for testosterone (nmol/L) to analyze the association between SHBG and cholecystectomy risk, and vice versa.

Potential nonlinear relationships between SHBG or testosterone and cholecystectomy risk were examined by applying restricted cubic splines with four knots at the 5th, 35th, 65th and 95th percentiles of the distribution of SHBG or testosterone in men or women. Subgroup analyses were performed to examine the relationships between SHBG or testosterone levels and cholecystectomy risk in men and in women, according to the following participant characteristics: age, ethnicity, smoking status, drinking status, diabetes, and BMI, in addition to menopausal status and HRT use in women.

In addition, to estimate the proportion of the obesity-cholecystectomy association that could be potentially mediated by the two sex hormones, we conducted mediation analyses using PROC CAUSALMED in SAS. The statistical tests conducted in this study were two-sided and the analyses were performed using Stata software (version 15.1; StataCorp) and SAS (version 9.4).

## Results

### Baseline Participant Characteristics

**Table 1** presents sex-specific characteristics of the study participants according to quartiles of SHBG levels in men and women. Regardless of sex, participants with higher levels of SHBG were more likely to be current smokers, had higher physical activity and lower BMI levels, and were less likely to have diabetes. Men with higher SHBG levels were older, whereas women with higher SHBG were younger. Women with higher SHBG levels were less likely to be menopausal or use HRT.

**Table 1.** Baseline participant characteristics according to quartile for Sex hormone-binding globulin (SHBG) in men and women.

|  | Quartile for SHBG in men (n = 176,909) |  |  |  | Quartile for SHBG in women (n = 160,147) |  |  |  |
| --- | --- | --- | --- | --- | --- | --- | --- | --- |
|  | Q1 | Q2 | Q3 | Q4 | Q1 | Q2 | Q3 | Q4 |
| Age, y | 53.0 ± 8.1 | 55.7 ± 8.1 | 57.4 ± 7.9 | 59.3 ± 7.4 | 56.1 ± 7.7 | 56.1 ± 8.0 | 55.6 ± 8.2 | 54.5 ± 8.3 |
| Ethnicity, % |  |  |  |  |  |  |  |  |
| White | 93.1 | 95.3 | 96.3 | 97.2 | 93.7 | 95.5 | 96.5 | 96.9 |
| Asian or Asian British | 4.4 | 2.4 | 1.8 | 1.3 | 3.3 | 1.8 | 1.2 | 0.9 |
| Black or Black British | 1.8 | 1.7 | 1.5 | 1.1 | 2.2 | 2.0 | 1.7 | 1.5 |
| Mixed | 0.7 | 0.5 | 0.5 | 0.4 | 0.7 | 0.7 | 0.7 | 0.7 |
| Townsend deprivation index | -1.12 ± 3.2 | -1.34 ± 3.1 | -1.37 ± 3.1 | -1.26 ± 3.2 | -1.15 ± 3.1 | -1.41 ± 3.0 | -1.48 ± 3.0 | -1.44 ± 3.0 |
| Smoking status, % |  |  |  |  |  |  |  |  |
| Never | 51.8 | 50.3 | 48.9 | 47.4 | 60.6 | 60.4 | 61.1 | 61.8 |
| Former | 37.8 | 38.4 | 38.4 | 37.0 | 31.4 | 30.7 | 30.0 | 28.7 |
| Current | 10.4 | 11.3 | 12.6 | 15.6 | 8.0 | 8.9 | 9.3 | 9.5 |
| Drinking status, % |  |  |  |  |  |  |  |  |
| Never | 3.4 | 2.6 | 2.4 | 2.7 | 7.3 | 5.4 | 4.9 | 5.1 |
| Former | 3.0 | 2.9 | 3.2 | 4.2 | 3.5 | 2.9 | 2.8 | 3.6 |
| Current | 93.6 | 94.5 | 94.5 | 93.1 | 89.3 | 91.7 | 92.3 | 91.3 |
| Physical activity, MET-h/week | 41.4 ± 46.6 | 45.1 ± 47.8 | 48.9 ± 50.2 | 52.5 ± 51.5 | 37.5 ± 39.2 | 40.8 ± 40.4 | 43.1 ± 40.9 | 45.2 ± 42.3 |
| Diabetes, % | 11.4 | 7.6 | 6.2 | 5.3 | 9.1 | 3.1 | 1.8 | 1.6 |
| BMI, kg/m<sup>2</sup> | 29.4 ± 4.3 | 28.3 ± 4.0 | 27.4 ± 3.9 | 26.2 ± 3.9 | 30.4 ± 5.5 | 27.6 ± 4.8 | 25.8 ± 4.3 | 24.4 ± 3.7 |
| Being menopausal, % | NA | NA | NA | NA | 74.2 | 71.4 | 67.4 | 59.7 |
| Ever used HRT, % | NA | NA | NA | NA | 35.2 | 34.5 | 33.2 | 33.1 |

BMI, body mass index; HRT, Hormone replacement therapy; MET, metabolic equivalent.

Median levels of SHBG were 38.88 nmol/L in men and 61.30 nmol/L in women, and median levels of testosterone were 11.92 nmol/L in men and 1.10 nmol/L in women. There was a positive correlation between SHBG and testosterone in men (r = 0.57) but not in women (r = –0.02) **(Supplementary** Figure 2). SHBG was moderately and inversely correlated with BMI both in men (r = –0.31) and in women (r = –0.47), whereas testosterone was inversely associated with BMI in men (r = –0.30) but not in women (r = 0.08).

### SHBG and Risk of Cholecystectomy

During a median follow-up of 12.27 years in men and 12.39 years in women, 2877 and 4607 incident cases of cholecystectomy were recorded in men and women, respectively.

After adjusting for sociodemographic factors, there were significant inverse associations between SHBG levels and risk of cholecystectomy, both in men and in women (**Table 2**). The associations in both sexes were attenuated but remained significant after further adjusting for lifestyle behaviors (model 2) and testosterone (model 3). The fully-adjusted HRs comparing the highest with the lowest quartiles of SHBG were 0.68 (95% CI: 0.59-0.77) in men (P-trend <0.001) and 0.39 (95% CI: 0.36-0.43) in women (P-trend <0.001). Each 1-SD increment in the SHBG levels was associated with a 13% (95% CI: 9%-18%) lower risk of cholecystectomy in men and a 31% (95% CI: 28%-33%) lower risk in women. The association appeared to be nonlinear in women (P-nonlinearity < 0.0001) but not in men (P-nonlinearity = 0.58) (**Figure 1**).

**Figure 1.**
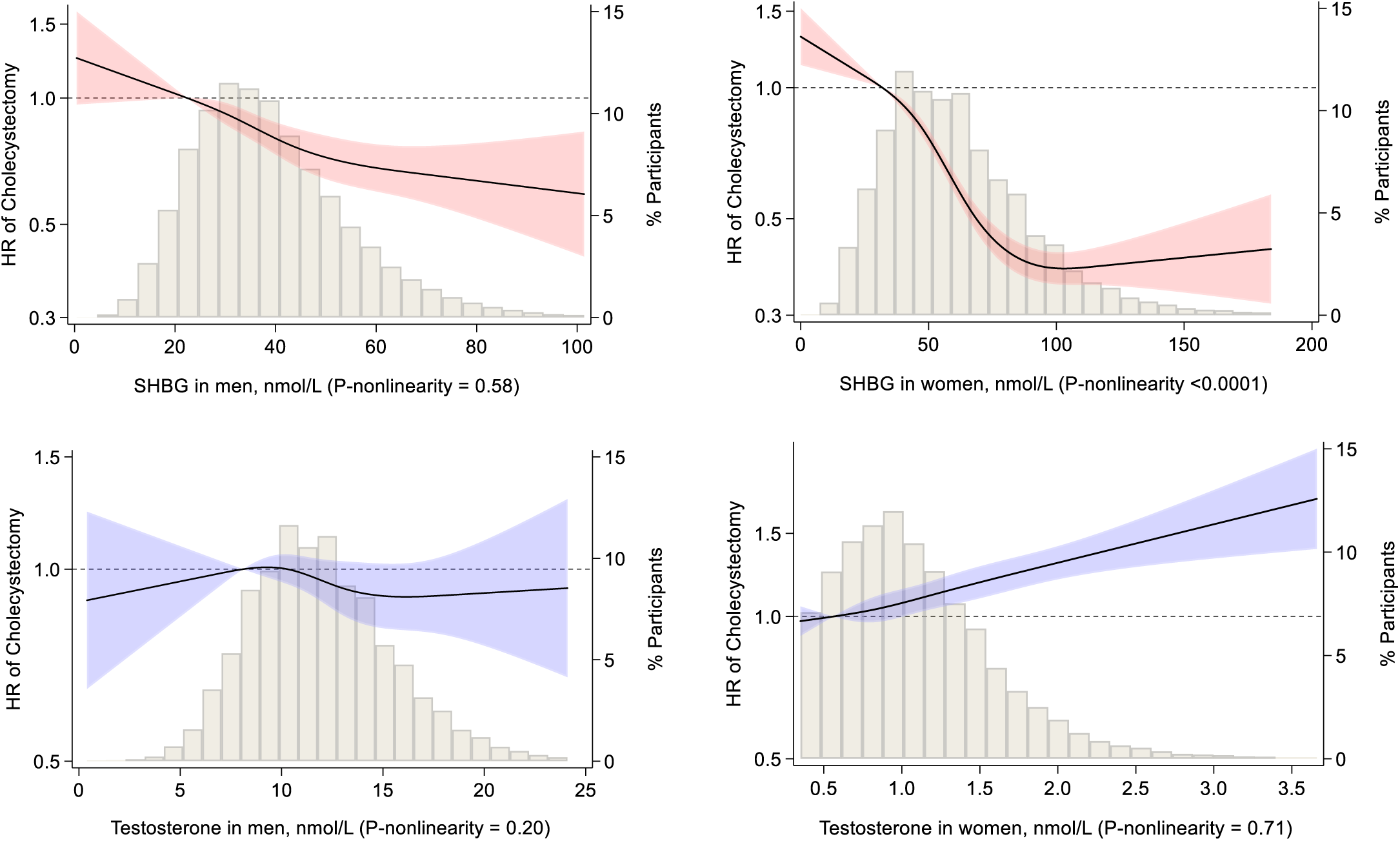
Restricted cubic splines for the associations of sex hormone-binding globulin (SHBG) or testosterone with risk of cholecystectomy in men and women. Median for the first quartile of SHBG or testosterone was used as the reference. Results were adjusted for covariates listed for model 3 in Table 2 or Table 3.

**Table 2.** Sex hormone-binding globulin (SHBG) and risk of cholecystectomy in men and women.

|  | Sex-specific quartile for SHBG |  |  |  | P-trend | Per SD increment |
| --- | --- | --- | --- | --- | --- | --- |
|  | Q1 | Q2 | Q3 | Q4 |  |  |
| Men |  |  |  |  |  |  |
| Median (range), nmol/L | 22.4 (<27.7) | 32.2 (27.7-36.6) | 41.5 (36.7-47.6) | 56.9 (≥47.7) |  |  |
| No. of participants | 44,195 | 44,252 | 44,207 | 44,255 |  |  |
| No. of cases/person-y | 803/547,379 | 751/545,427 | 704/541,876 | 619/536,620 |  |  |
| Model 1 (HR [95% CI]) | 1.00 (referent) | 0.86 (0.78-0.95) | 0.76 (0.69-0.85) | 0.64 (0.57-0.71) | <0.001 | 0.85 (0.81-0.88) |
| Model 2 (HR [95% CI]) | 1.00 (referent) | 0.87 (0.79-0.96) | 0.78 (0.70-0.86) | 0.64 (0.57-0.72) | <0.001 | 0.85 (0.82-0.88) |
| Model 3 (HR [95% CI]) | 1.00 (referent) | 0.88 (0.80-0.98) | 0.80 (0.72-0.90) | 0.68 (0.59-0.77) | <0.001 | 0.87 (0.82-0.91) |
| Women |  |  |  |  |  |  |
| Median (range), nmol/L | 31.5 (<40.5) | 48.5 (40.5-56.7) | 65.7 (56.8-77.7) | 94.2 (≥76.8) |  |  |
| No. of participants | 40,017 | 40,047 | 40,037 | 40,046 |  |  |
| No. of cases/person-y | 1745/491,357 | 1309/495,327 | 876/498,581 | 677/498,953 |  |  |
| Model 1 (HR [95% CI]) | 1.00 (referent) | 0.75 (0.69-0.80) | 0.49 (0.45-0.54) | 0.38 (0.35-0.42) | <0.001 | 0.68 (0.65-0.70) |
| Model 2 (HR [95% CI]) | 1.00 (referent) | 0.77 (0.72-0.83) | 0.52 (0.48-0.56) | 0.39 (0.36-0.43) | <0.001 | 0.69 (0.67-0.71) |
| Model 3 (HR [95% CI]) | 1.00 (referent) | 0.77 (0.72-0.83) | 0.52 (0.48-0.56) | 0.39 (0.36-0.43) | <0.001 | 0.69 (0.67-0.72) |

Model 1: age (y), ethnic group (White, Asian/Asian British, Black/Black British, mixed), and Townsend deprivation index.
Model 2: model 1 + smoking (never, former, current [ $<10$ , $10-<50$ , $\geq 50$ pack-years]), alcohol drinking (never, former, current $<10$ g/d in men or $<5$ g/d in women, current $10-<25$ g/d in men or $5-<15$ g/d in women, current $\geq 25$ g/d in men or $\geq 15$ g/d in women), total physical activity (MET-h/week), diabetes (yes, no), and for women, menopausal status (yes, no) and hormone replacement therapy (ever, never).
Model 3: model 2 + testosterone (nmol/L).

**Table 3.** Testosterone and risk of cholecystectomy in men and women.

|  | Sex-specific quartile for testosterone |  |  |  | P-trend | Per SD increment |
| --- | --- | --- | --- | --- | --- | --- |
|  | Q1 | Q2 | Q3 | Q4 |  |  |
| Men |  |  |  |  |  |  |
| Median (range), nmol/L | 8.08 (<9.44) | 10.5 (9.44-11.6) | 12.7 (11.6-14.1) | 16.0 (≥ 14.1) |  |  |
| No. of participants | 44,215 | 44,226 | 44,236 | 44,232 |  |  |
| No. of cases/person-y | 842/539,332 | 767/543,266 | 678/544,777 | 590/543,926 |  |  |
| Model 1 (HR [95% CI]) | 1.00 (referent) | 0.92 (0.83-1.01) | 0.81 (0.73-0.90) | 0.72 (0.65-0.80) | <0.001 | 0.88 (0.85-0.92) |
| Model 2 (HR [95% CI]) | 1.00 (referent) | 0.94 (0.85-1.04) | 0.84 (0.76-0.93) | 0.74 (0.66-0.82) | <0.001 | 0.89 (0.86-0.93) |
| Model 3 (HR [95% CI]) | 1.00 (referent) | 1.00 (0.90-1.11) | 0.94 (0.85-1.05) | 0.91 (0.80-1.04) | 0.12 | 0..97 (0.92-1.02) |
| Women |  |  |  |  |  |  |
| Median (range), nmol/L | 0.56 (<0.72) | 0.87 (0.72-1.02) | 1.17 (1.02-1.36) | 1.67 (≥ 1.37) |  |  |
| No. of participants | 39,911 | 40,113 | 40,065 | 40,058 |  |  |
| No. of cases/person-y | 1058/494,210 | 1126/497,376 | 1092/497,136 | 1331/495,496 |  |  |
| Model 1 (HR [95% CI]) | 1.00 (referent) | 1.06 (0.98-1.15) | 1.03 (0.95-1.13) | 1.26 (1.16-1.37) | <0.001 | 1.10 (1.07-1.13) |
| Model 2 (HR [95% CI]) | 1.00 (referent) | 1.08 (0.99-1.18) | 1.07 (0.98-1.16) | 1.32 (1.21-1.43) | <0.001 | 1.11 (1.08-1.14) |
| Model 3 (HR [95% CI]) | 1.00 (referent) | 1.07 (0.99-1.17) | 1.06 (0.97-1.15) | 1.28 (1.18-1.39) | <0.001 | 1.10 (1.07-1.13) |

Model 1: age (y), ethnic group (White, Asian/Asian British, Black/Black British, mixed), and Townsend deprivation index.
Model 2: model 1 + smoking (never, former, current [ $<10$ , $10-<50$ , $\geq 50$ pack-years]), alcohol drinking (never, former, current $<10$ g/d in men or $<5$ g/d in women, current $10-<25$ g/d in men or $5-<15$ g/d in women, current $\geq 25$ g/d in men or $\geq 15$ g/d in women), total physical activity (MET-h/week), diabetes (yes, no), and for women, menopausal status (yes, no) and hormone replacement therapy (ever, never).
Model 3: model 2 + SHBG (nmol/L).

### Testosterone and Risk of Cholecystectomy

After the adjustment for sociodemographic factors, testosterone was significantly and inversely associated with risk of cholecystectomy in men (P-trend <0.001), but was positively associated with the risk in women (P-trend <0.001) (**Table 3**). In men, the association remained significant after further adjusting for lifestyle behaviors (model 2). However, after an additional adjustment for SHBG (model 3), there was no association between testosterone and risk of cholecystectomy in men (HR_Q4 vs. Q1_ = 0.91, 95% CI: 0.80-1.04; P-trend = 0.12).

In women, the positive association between testosterone and risk of cholecystectomy persisted in the fully adjusted model (model 3), with an HR of 1.28 (95% CI: 1.18-1.39; P trend <0.001) comparing the highest with the lowest quartiles. Each 1-SD increment in testosterone levels was associated with a 10% (95% CI: 7%-13%) higher risk of cholecystectomy. There was no evidence for a nonlinear association between testosterone and cholecystectomy risk either in men or in women (Figure 1).

### Subgroup Analysis

In men, the inverse association between SHBG levels and risk of cholecystectomy was broadly consistent across various subgroups defined by demographic characteristics and other risk factors (**Figure 2**). There was evidence that the association was attenuated with increasing BMI (P-interaction = 0.044). In women, there was evidence for a stronger association among those who did not smoke (P-interaction <0.001), never drink (P-interaction = 0.001), or never used HRT (P-interaction <0.001) than the association in the corresponding comparison groups. For testosterone, the association with cholecystectomy in men was inverse in a few subgroups (non-white ethnicity, former drinkers, and normal-BMI group), whereas the association in women appeared to be stronger in current drinkers than in never or former drinkers without other group differences.

**Figure 2.**
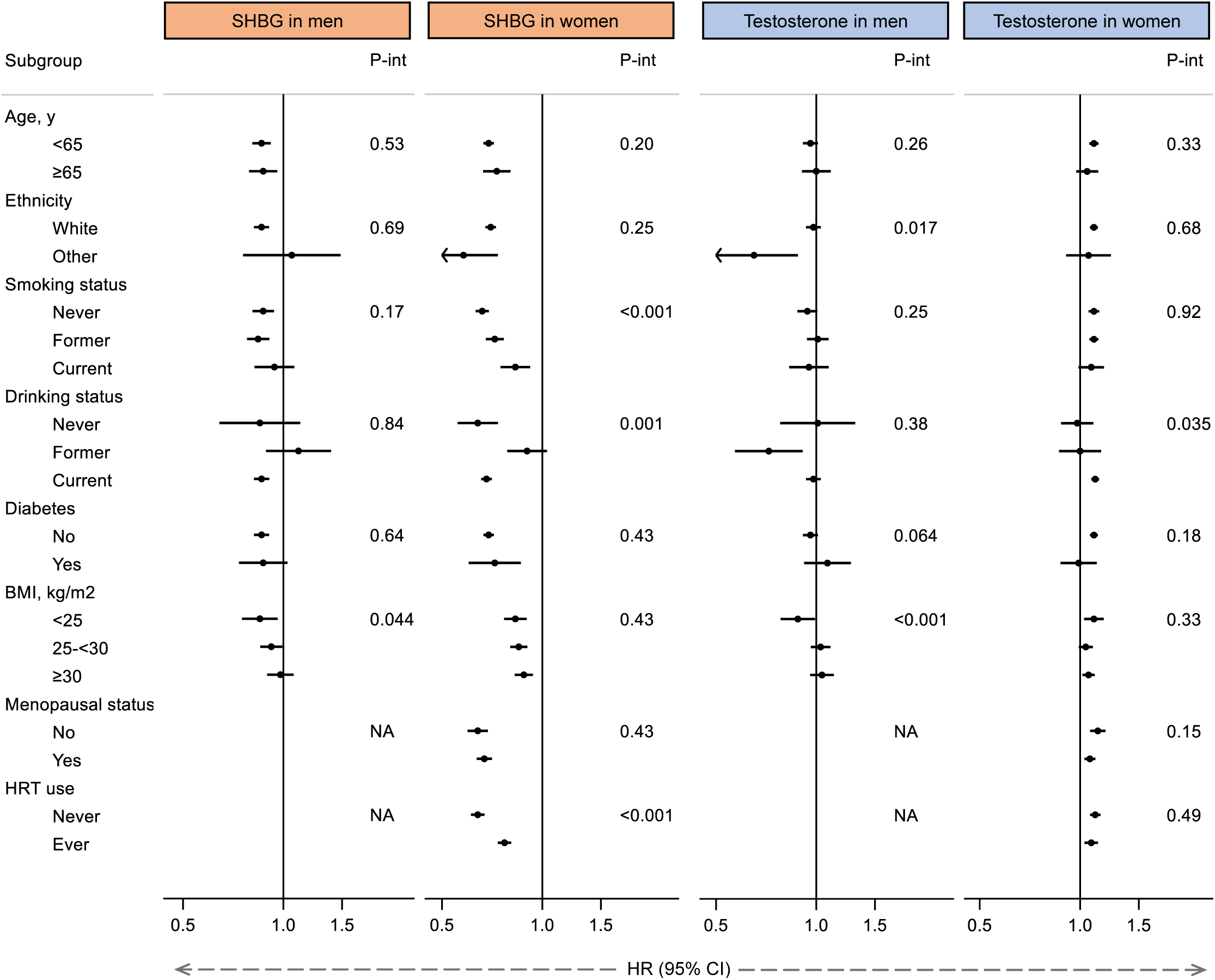
Subgroup analysis for the associations of sex hormone-binding globulin (SHBG) or testosterone with risk of cholecystectomy in men and women. Results were HR (95% CI) of cholecystectomy associated with per SD increment in SHBG or testosterone after adjustment for the covariates listed for model 3 in Table 3.

### Mediation Analysis

After multivariable adjustment, being obesity (BMI ≥ 30 vs. <30 kg/m^2^) was associated with a 148% (HR = 2.48, 95% CI: 2.32-2.63) higher risk of cholecystectomy in women and a 63% (HR = 1.63, 95% CI: 1.50-1.76) higher risk in men. For women, results from the mediation analysis indicated that 14.71% (95% CI: 8.23%-21.19%) and 2.74% (95% CI: 1.37%-4.11%) of the obesity-cholecystectomy association was mediated by serum levels of SHBG and testosterone, respectively (**Figure 3**). No significant medication was observed for either hormone in men (data not shown).

**Figure 3.**
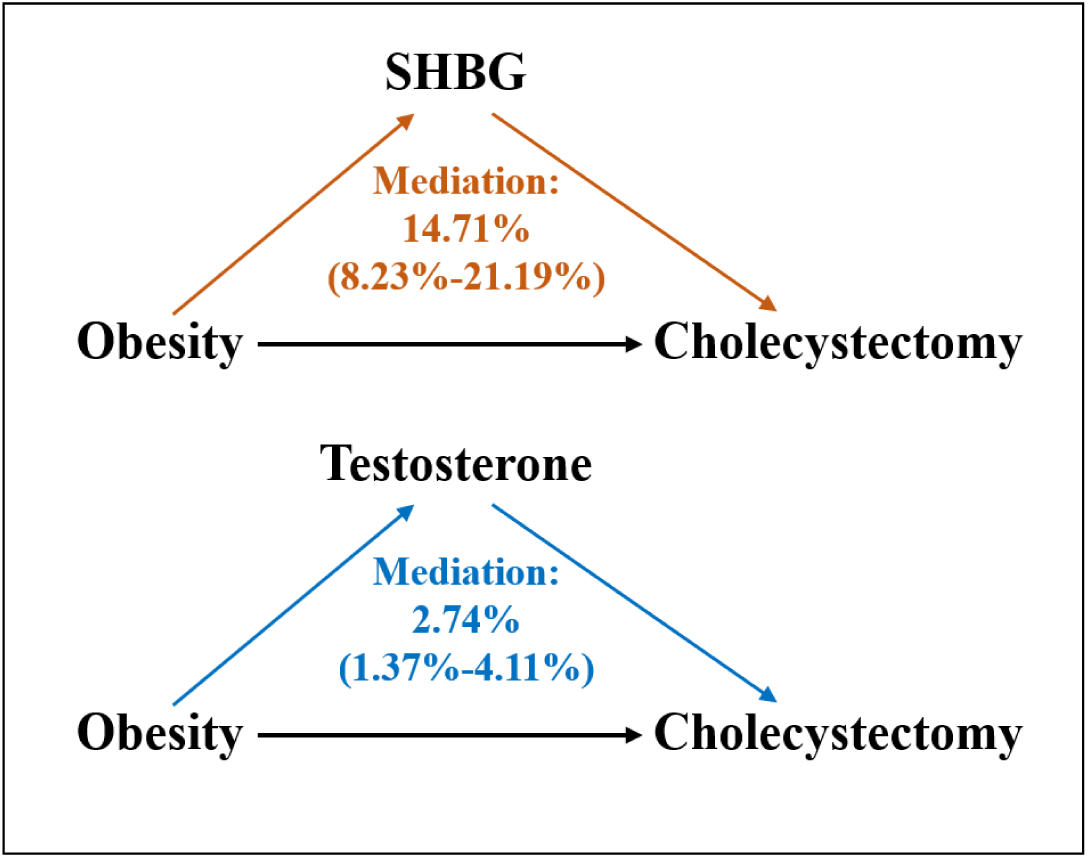
Proportion of the association between obesity and risk of cholecystectomy potentially mediated by sex hormone-binding globulin (SHBG) or testosterone in women.

## Discussion

In this prospective analysis of middle-aged and older adults in UK, we systematically assessed two sex hormones for their relationships with risk of cholecystectomy in men and women. Higher levels of SHBG were associated with a lower risk of cholecystectomy in both sexes. Higher levels of testosterone were associated with a higher risk of cholecystectomy in women but not in men. In addition, we found that obesity showed sex-specific associations with risk of cholecystectomy, with a substantially stronger association in women than in men, which is in line with previous evidence from a Mendelian randomization (MR) study (19). We further estimated that 14.71% and 2.74% of the association between obesity and cholecystectomy was significantly mediated by serum levels of SHBG and testosterone, respectively, whereas no mediation was observed for neither hormone in men.

Very few previous studies have assessed circulating levels of SHBG in relation to risk of gallstone diseases. In a 1:1 matched case-control study from Italy, which included 84 pairs of men and women aged 30-69 years, SHBG levels did not differ between cases and controls, regardless of sex (16). In a cohort study of the general population in Denmark, Shabanzadeh et al. (17) assessed SHBG levels in association with incident gallstones in 1227 men aged 30-60 years. They found that baseline levels of SHBG were inversely associated with incident gallstones, but the association disappeared with an additional adjustment for BMI (17), which represent a potential overall adjustment.

However, longitudinal increases in SHBG were found to be inversely associated with incident gallstones after multivariable adjustment including adjustment for baseline and changes in BMI (17). In the present study with a substantially larger number of outcome events, we found a consistent inverse association between serum level of SHBG and risk of cholecystectomy both in men and in women. In women, SHBG mediated 14.71% of the association between obesity and cholecystectomy, suggesting SHBG as a biomarker for cholecystectomy and the potential therapeutic target for gallbladder disease.

We found that testosterone levels were positively associated with cholecystectomy risk in women but not in men, independently of other covariates including SHBG. Animal studies have shown that when fed the lithogenic diet, no cholesterol gallstones formed in any of the female hamsters (24). However, upon the addition of methyltestosterone to the diet, the incidence of cholesterol gallstones increased from 0% to 86% after 6 weeks (13), indicating the role of testosterone in the process of gallstone formation. Previous studies of testosterone and incident gallstones have reported inconsistent results (15–17). The Italian case-control study found no association between testosterone levels and cholesterol gallstones (16). In another case-control study including relatively younger (<50 years old) men and women, compared to a control group of hospitalized patients without stones, men with symptomatic radiolucent gallstones showed lower testosterone levels (15). In the Danish cohort study by Shabanzadeh et al. (17), neither baseline nor longitudinal changes in total testosterone were associated with incident gallstones.

However, higher levels of free testosterone at baseline (but not the longitudinal changes) were significantly and positively associated with incident gallstones (17). In line with our findings, a MR study observed that genetically-determined higher levels of testosterone were associated with a higher risk of cholelithiasis in women (18). In the present study, we also observed that testosterone mediated a small proportion of the association between obesity and cholecystectomy in women. These findings collectively support testosterone as an intervention target for reducing the risk of cholelithiasis in women.

The underlying mechanisms through which SHBG and testosterone might influence the risk of cholecystectomy have not been well understood. SHBG is predominantly synthesized by the liver and is released into the bloodstream, where it strongly attaches to sex hormones, influencing their bioavailability (25). However, it is still unclear whether SHBG directly influences the underlying processes of gallstone diseases or affects them indirectly by controlling the access of sex hormones to tissues. Impaired gallbladder motility is an important contributing factor in cholesterol gallstone disease (26). Kline et al. (27) found that testosterone and its active metabolite rapidly inhibits gallbladder motility in guinea pigs through nongenomic hormone actions. These effects encompass pathways such as the inhibition of intracellular Ca2+ release, the hindrance of extracellular Ca2+ entry, and the involvement of PKC. Another possible pathological mechanism involves cholesterol absorption in the liver. Testosterone deficiency triggers pronounced hypercholesterolemia, and markedly reduced hepatic LDLR mRNA expression and protein levels, leading to attenuated LDL cholesterol clearance (28). This can be reversed through testosterone replacement therapy, resulting in improved cholesterol absorption in the liver and elevated bile concentration (28, 29).

The present study is the first prospective study to systematically investigate the sex-specific associations of serum levels of SHBG and testosterone with risk of cholecystectomy. The study utilized a large sample size with long-term follow up and an adequate number of outcome events. We also extensively considered a wide arrange of potential confounding factors and incorporated them into the multivariable models to ensure the reliability and authenticity of the findings. In addition, our findings on the mediation roles of serum hormones in the impact of obesity on cholecystectomy in women may provide novel mechanistic insight into the sex-specific difference in the association between obesity and cholelithiasis.

There are some limitations to be acknowledged. First, due to the observational nature of the study, causality cannot be established. Despite the careful adjustment for a variety of known risk factors, we could not completely exclude the potential influence of residual confounding on our results. Second, baseline participant characteristics such as drinking status, smoking status, and diet were collected via self-reported touch-screen questionnaires, which may have introduced recall bias. Third, the assessed sex hormones were measured at baseline without accounting for changes in the hormone levels over time. Fourth, for testosterone, only total levels but not free testosterone levels, which may be more closely associated with gallbladder disease (17), were measured in the UK Biobank. Finally, our analysis included participants of European descent, who have a higher socioeconomic status and are relatively healthier as compared with the general adult population in the UK (30). Therefore, our findings need to be confirmed for other regional and ethnic populations.

## Conclusions

In summary, our findings suggest that SHBG levels are inversely associated with risk of cholecystectomy irrespective of sex, whereas testosterone levels are positively associated with risk of cholecystectomy only in women. Serum SHBG and testosterone significantly mediated the relationship between obesity and cholecystectomy in women but not in men. These findings support the involvement of sex hormones in the development of gallbladder disease, and may provide some novel insights into the sex differences in the risk factors for and incidence of cholelithiasis.

## Data Availability

UK Biobank data can be requested by bona fide researchers for approved projects, including replication, through https://www.ukbiobank.ac.uk/.

## Acknowledgements

The authors thank the UK Biobank participants. This research was conducted using the UK Biobank Resource under application number 90087.

## Abbreviations

BMI, body mass index; CI, confidential interval; HbA1c, hemoglobin A1c; HR, hazard ratio; HRT, replacement therapy; ICD, International Classification of Disease; MET, metabolic equivalent; SD, standard deviation; SHBG, sex hormone-binding globulin.

## Author contributions

G-CC designed the research and developed the analytical plan. J-QL performed the statistical analyses and had primary responsibility for writing the manuscript. G-CC and Y-FH directed the study. All authors contributed to the interpretation of data, critically reviewed and revised the manuscript, and approved the final manuscript. G-CC is the guarantor of this work and, as such, had full access to all the data in the study and takes responsibility for the integrity of the data and the accuracy of the data analysis.

## Funding

No funding.

## Declarations

**Ethics approval and consent to participate:** was approved by the UK North West Multicenter Research Ethics Committee, and all participants provided informed written consent.

## Consent for publication

Not applicable.

## Competing interests

The authors declare that they have no competing interest.

## Notes

### Competing Interest Statement

The authors have declared no competing interest.

### Funding Statement

This study did not receive any funding

