## Supplementary Figure 1, Supplementary Figure 2 for "Sex hormones, obesity, and risk of cholecystectomy in men and women: a population-based prospective study and mediation analysis"

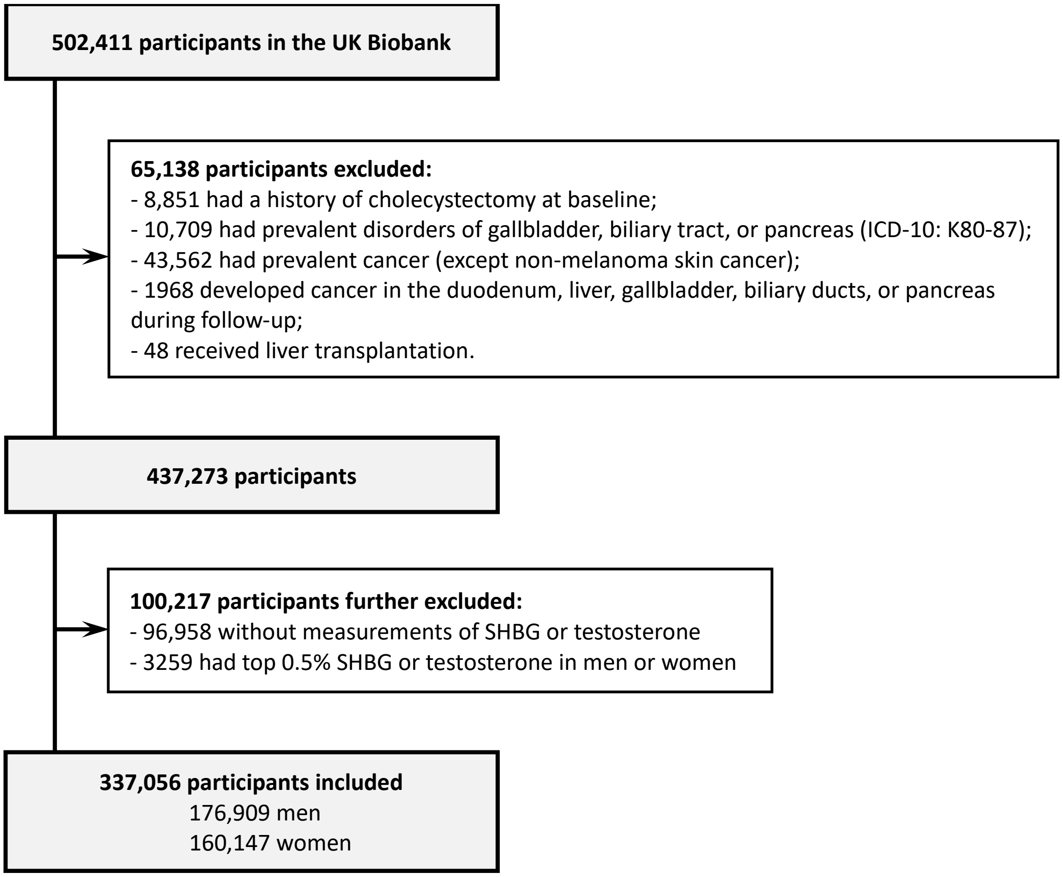


**Supplementary Figure 1. Participant selection.**

**Men**

| | SHBG | Testosterone | Age | BMI |
|---|---|---|---|---|
| SHBG | 1.00 | 0.57 | 0.29 | -0.31 |
| Testosterone | 0.57 | 1.00 | -0.06 | -0.30 |
| Age | 0.29 | -0.06 | 1.00 | 0.03 |
| BMI | -0.31 | -0.30 | 0.03 | 1.00 |

**Women**

| | SHBG | Testosterone | Age | BMI |
|---|---|---|---|---|
| SHBG | 1.00 | -0.02 | -0.07 | -0.47 |
| Testosterone | -0.02 | 1.00 | -0.14 | 0.08 |
| Age | -0.07 | -0.14 | 1.00 | 0.10 |
| BMI | -0.47 | 0.08 | 0.10 | 1.00 |

**Supplementary Figure 2. Spearman correlation among SHGB, testosterone, age, and BMI.**
